# Empirical phenotyping of joint patient-care data supports hypothesis-driven investigation of mechanical ventilation consequences

**DOI:** 10.1101/2023.12.14.23299978

**Authors:** J.N. Stroh, Peter D. Sottile, Yanran Wang, Bradford J. Smith, Tellen D. Bennett, Marc Moss, David J. Albers

## Abstract

Analyzing patient data under current mechanical ventilation (MV) management processes is essential to understand MV consequences over time and to hypothesize improvements to care. However, progress is complicated by the complexity of lung-ventilator system (LVS) interactions, patient-care and patient-ventilator heterogeneity, and a lack of classification schemes for observable behavior. Ventilator waveform data originate from patient-ventilator interactions within the LVS while care processes manage both patients and ventilator settings. This study develops a computational pipeline to segment joint waveform and care settings timeseries data into phenotypes of the data generating process. The modular framework supports many methodological choices for representing waveform data and unsupervised clustering. The pipeline is generalizable although empirical output is data- and algorithm-dependent. Applied individually to 35 ARDS patients including 8 with COVID-19, a median of 8 phenotypes capture 97% of data using naive similarity assumptions on waveform and MV settings data. Individual’s phenotypes organize around ventilator mode, PEEP, and tidal volume with additional delineation of waveform behaviors. However, dynamics are not solely driven by setting changes. Fewer than 10% of phenotype changes link to ventilator settings directly. Evaluation of phenotype heterogeneity reveals LVS dynamics that cannot be discretized into sub-phenotypes without additional data or alternate assumptions. Individual phenotypes may also be aggregated for use in scalable analysis, as behaviors in the 35 patient cohort comprise 16 cohort-scale LVS types. Further, output phenotypes compactly discretize the data for longitudinal analysis and may be optimized to resolve features of interest for specific applications.

## 1 Introduction

Mechanical ventilation (MV) of critical care patients provides life-saving support over periods typically lasting days to weeks. Over these timescales, ventilator management strategies significantly impact patient outcome^1,2^. Modern care protocols and technologies^3,4^ emphasize lung-protective strategies^5^ to minimize potentially deleterious consequences of MV. These include ventilator-induced lung injury (VILI,^6^) and patient-ventilator dyssynchrony (PVD), a disagreement between ventilator action and patient effort. Both PVD and VILI may contribute to acute respiratory distress syndrome (ARDS) and ARDS-related mortality^7–9^. Protective strategies depend on mechanistic understanding to guide ventilator settings, including positive end-expiratory pressure (PEEP), tidal volume, and driving pressure^10–12^. Despite the effectiveness of protective advances, association between MV and ARDS-related mortality remains unacceptably high. Reducing these highly negative outcomes motivates continued improvement and personalization of VILI-minimizing ventilator strategies^13–15^.

Hypotheses about current care are essential for improving MV, but such scientific inquiry suffers from a lack of general breath categories and understanding of patient-ventilator variability. MV applied in critical care typically lasts 3–7 days^16–18^ amidst the context of non-stationary patient conditions and other care procedures. While the short-duration physiology under MV is understood, analysis of those relationships and MV consequences over longer therapeutic timescales are limited and hindered by patient- and care-specific heterogeneity. A method for labeling and classifying MV breaths based on characteristics is desirable to reduce data complexity and facilitate temporal analysis. Currently, the most accessible classification scheme identifies PVD types from waveform characteristics. PVD research is rich with ML applications primarily focused on extending manual labels to larger datasets through supervised methods^19–22^, identifying PVD waveform characteristics^23^, and estimating event severity^24^. However, these labels may be ill-suited for MV research involving temporal analysis: they are stationary, are not mutually exclusive^21^, depend on MV mode characteristics^8^, and vary in organization^25,26^. Another research avenue uses interpretable model-based parametrizations to analyze waveform data^27–31^, potentially allowing for a wider and more flexbile exploration of breath behavior.

The clinical observables from MV include airway pressure (*p*), volume (*V* ), and flow timeseries that record the dynamic interaction between patient lungs and care-managed machine. The human lung-ventilator system (LVS), rather than an isolated human lung, underlies the data generating process when investigating MV from waveform-sourced data^29^. Moreover, MV management changes ventilator settings; these care factors contextualize the waveform data within LVS trajectories. The assemblages of coupled LVSs and applied management processes are the data generating process and the objects of interest for improving MV.

Quantifying clinical consequences of MV on patient health are necessary to evaluate and improve MV care. This requires linking outcomes to MV descriptors that include both ventilator setting as well as patient-ventilator interaction, but these LVS categories do not exist. Analysis based on ventilator settings and care processes alone will not incorporate patient-heterogeneous responses observed in pressure-volume, while those based on PVD labels that omit ventilator settings. This work addresses the methodological gap in quantifying MV consequences by developing an unsupervised categorization process to define joint LVS state categories as suitable targets for consequence association. Namely, it digitizes joint LVS data into interpretable phenotypes based on data similarities^32^. This approach reduces the dimensionality of the problem, enabling scalability to larger datasets, while incorporating the essential data components needed for consequence attribution.

Clinical validation of phenotypes requires linking breath behaviors to outcomes or other MV consequences. This work develops a generalized process to produce validatable phenotypes with clinical validation an intended downstream application (Sec4).

In this work, phenotype trajectories of individuals are scrutinized in relation to MV management changes and timeseries of PVD labels to evaluate their consistency and ability to differentiate important characteristics. The phenotyping examples assign equal weight to LVS feature components to be agnostic about data element importance. The resulting data segmentation follow ventilator settings changes and persistent variations in waveform behavior within individuals individuals timeseries, while aggregate cohort-scale analysis shows they are general enough to mix patients. The main result is a generalized phenotyping pipeline whose empirical results are data-specific phenotypes. These outputs are not anticipated not generalize beyond the 2-day snippets of 35 ARDS patients on one ventilator model, because the data do not represent the broad diversity of MV breaths. The data-specific classification approach is tied to context of the data, providing benefits of informativeness and accuracy generally lost in a universal scheme^33^.

A robust and systematic process for phenotyping the diverse LVS behaviors is a necessary step toward quantifying MV consequences and optimization of respiratory management to mitigate VILI. Phenotypes output by the developed pipeline may be used as a basis for explaining impacts on respiratory health. For example, one could investigate the distribution of phenotype occurrences, combining both ventilator settings and patient-ventilator response to them, with temporal changes in driving pressure^34^ or gas ratios^35,36^. Importantly, phenotypes in this context data mask low-level heterogeneity to reduce trajectory complexity (§3.1) and provide a standard basis for comparison across patients (§3.2).

## 2 Method

Phenotype identification analyzes LVS data, including waveforms and ventilator settings, using an unsupervised computational pipeline. This section develops a specific implementation while framing it in a general way. The process is generalized but its output may not be: *empirical phenotypes reflect the data and methods defining them*.

The modular workflow enables adjustments to the source data, waveform representation, feature definition, and segmentation strategy. This permits phenotype generation to accommodate different hypotheses about which aspects of the patient-ventilator-care system matter when evaluating MV consequence or other targets.

### 2.1 Data

Data including airway pressure, volume, and ventilator settings were captured for a cohort of intubated at University of Colorado ICUs with ARDS diagnoses, were mechanical ventilated using Hamilton G5 ventilators (https://www.hamilton-medical.com), and who had substantial risk of VILI. The collection effort and data use were approved by Colorado Multiple Institutional Review Board protocol (COMIRB, protocol #18-1433) and follow ethical standards set by COMIRB and the Helsinki Declaration of 1975. Children, pregnant women, and age-censored elders (*>* 89 years), and the imprisoned were excluded. Enrollment targeted collection of esophageal pressures, which are not analyzed in this work, and imposed additional exclusion criteria (*viz*. esophageal fistula, variceal bleeding or banding, facial fracture, and recent gastric/esophageal surgery). Eligibility for recording was contingent on active MV therapy, so patients were necessarily unconscious at the time of enrollment. Each patient’s identified proxy decision-maker provided informed consent as unconscious patients could not consent directly.

Following esophageal balloon placement, continuous recordings up to 48 hours were made directly from ventilators for 35 MV encounters satisfying enrollment criteria^22^. The cohort includes 14 women and 21 men with median age 58 years and interquartile range (IQR) 24.9 years; 71.4% are white, 34% of which identify as Hispanic or Latino. Pre-processing comprised removal of breaths with ventilator calibration artifacts and carrying forward last-known settings values within ventilator modes. Table 1 summarizes clinical and demographic characteristics of included patients. Data total 1.74 million breaths over 71.14 recording-days (median 1.97[1.56] days per patient) recorded at 31.25 Hz. Adaptive pressure volume-controlled and pressure-controlled mandatory ventilation modes (APVCMV and P-CMV, respectively) account for 84% and 10% of breaths, respectively, with the remainder in spontaneous/supported (SPONT), synchronized controlled (SCMV), and standby modes. Care and ventilator management follow the ARDSnet protocols^7^.

**Table 1.** Tabular summary of the patient cohort and associated data. ‘Monitored’ and ‘Recorded’ denote the duration spanned by data and length continuous data contents, respectively, in hours. P:F ratio is the PaO_2_/FiO_2_ ratio at admission used to qualify ARDS and need for MV, AA = African-American, AI = American-Indian, AK = Alaska, NMB = Neuromuscular blockade

| Detail | Count | % | Median | IQR |
| --- | --- | --- | --- | --- |
| Monitored (hrs) |  |  | 47.1 | 38.2 |
| Recorded (hrs) |  |  | 43.7 | 39 |
| Age (years) |  |  | 58.0 | 24.8 |
| Gender |  |  |  |  |
| Female | 14 | 40 | 54.5 | 25.0 |
| Male | 21 | 60 | 58.0 | 26.5 |
| Race/Ethnicity |  |  |  |  |
| White | 25 | 71.4 |  |  |
| Unknown/NA | 5 | 14.3 |  |  |
| Black/AA | 3 | 8.6 |  |  |
| AI or AK Native | 1 | 2.9 |  |  |
| More than one race | 1 | 2.9 |  |  |
| ARDS risk |  |  |  |  |
| Pneumonia | 11 | 31.4 |  |  |
| COVID | 11 | 31.4 |  |  |
| Sepsis | 6 | 17.1 |  |  |
| Other | 3 | 8.6 |  |  |
| Pancreatitis | 2 | 5.7 |  |  |
| Aspiration | 2 | 5.7 |  |  |
| P:F ratio |  |  | 136.0 | 77.0 |
| Mortality | 8 | 22.9 |  |  |
| NMB use | 9 | 25.7 |  |  |

#### Dyssynchrony labels

Breath-wise PVD identified by supervised ML in previous work^19,22^ are used to enrich LVS evolution context and provide comparison for extracted categories. PVD types were assigned breath-wise to the data using a gradient boosted decision tree (XGBoost) based on a manually labeled subset. PVD categories include normal (NL), reverse triggered (RT), early flow limited (eFL), double trigger (DT), and early vent termination (EVT) types as defined and applied in previous analysis^21,37^. One-minute moving averages of these breath labels communicate PVD occurrence over time.

### 2.2 Pipeline

The computational pipeline described is a process for developing phenotypes from joint waveform and MV care-related data. The method is depicted in Figure 1 and follows Wang *at al*.^32^ by using data-informed parameter distributions to uncover latent similarities in observed ICU patients. The three main phases (feature construction, segmentation, and interpretation) involve methodological decisions, which are discussed below both in general terms and in terms of specific implementation applied to individual patient data. Code developed in MATLAB^®^ for the implemented pipeline is available by reasonable request to the authors.

**Figure 1.**
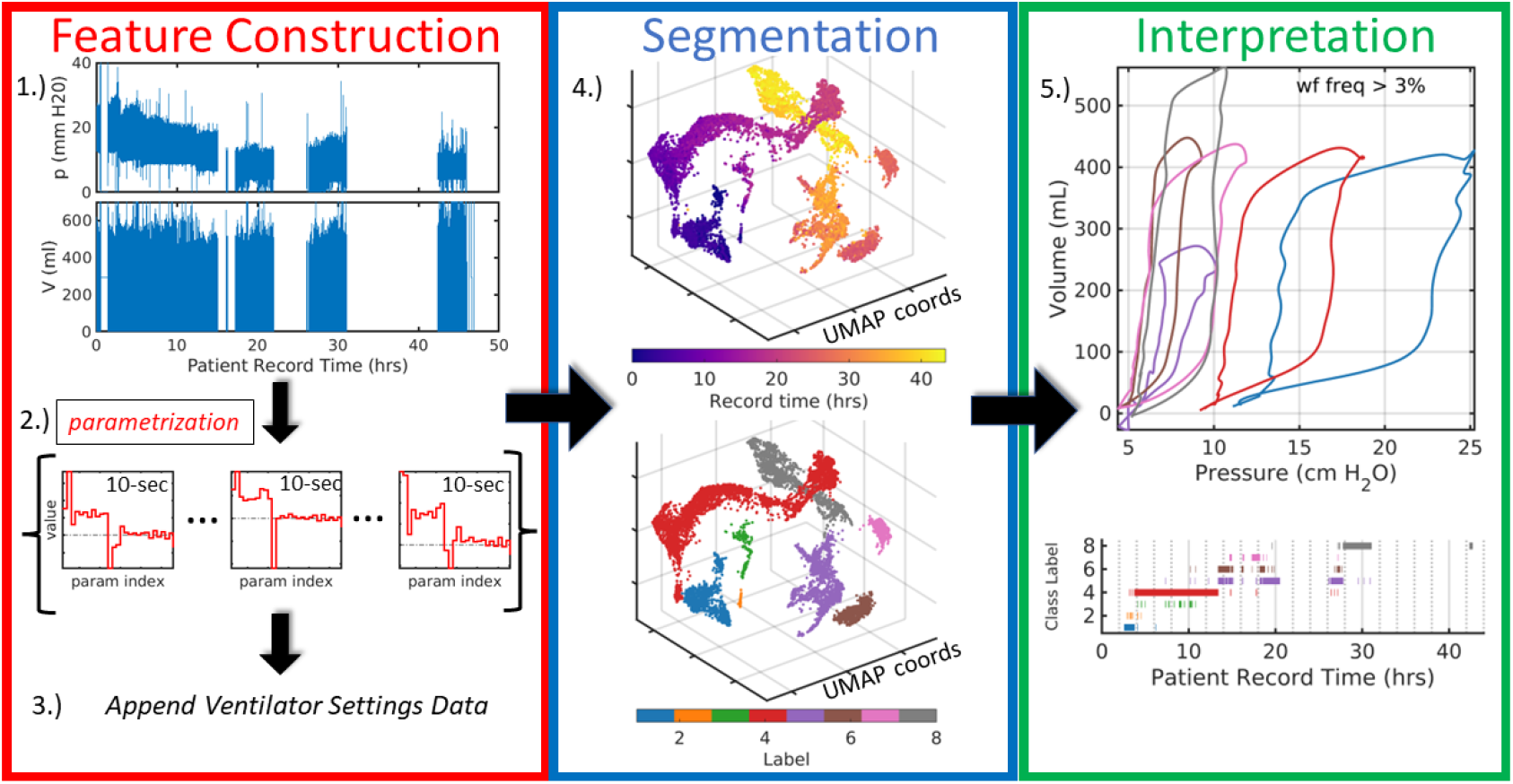
Broad pipeline organization. Raw data (1.) are digitally parametrized (2.) over short windows, typically satisfying stationarity assumptions. Distributional parameter estimates are summarized and augmented with the contextual data of ventilator settings (3.) which include information such as ventilator operation mode, positive-end expiratory pressure (PEEP) or other baseline pressure, flow and pressure triggers, and minimum mandatory breath rate. Feature vectors, defined by the augmented LVS descriptors, are reduced to three dimensions (4.) where they can be analyzed based on time ordering (top) and structural similarity via segmentation (bottom). Finally, in (5.), temporal evolution of the system is compactly encoded in the time-ordered LVS descriptor labels and their associated waveform characterizations in an interpretable and explainable way. The process transforms raw data (1.) into a more easily comprehensible form (5.).

#### 1.) Waveform parametrization

Waveform observations can optionally be digitized to improve comparison of breath-level data through approximation and regularization of the continuous time signals. Digitization examples in other work include spectrograms^38^, clinical parameters^19^, model parametrization^31^, or signal processing methods such as polynomials and wavelets.

One empirical parametrization developed for LVS data analysis^29^ uses an asynchronous ensemble Kalman smoother (wEnKS)^39^ to transform waveform data segments into parameter distributions (SI A). Specifically, observations *y*^*o*^ (representing pressure or volume data) are mapped to *M*-dimensional parameter vector samples {**a**} of the bayesian posterior distribution *p*(**a** | *y*^*o*^). The likelihood function *p*(*y*^*o*^ | **a**) describes the RMS error between data *y*^*o*^ and simulated counterparts defined by the ordinary differential equation

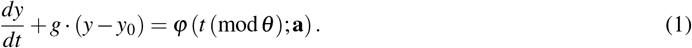

Here, *g* is a fixed smoothing parameter, *θ* is constant breath rate over the interval so that *t* (mod *θ* ) gauges time within each breath cycle, and *y*_0_ is the baseline value of the signal (PEEP when *y* is pressure). The function *ϕ* in Eq.1 is a piecewise constant function over the breath cycle with heights determined by parameter vector **a**, whose optimal values digitize the observational waveform data (details found in SI A.1 and past work^29^).

The model construction ensures parameter identification throughout a bayesian ensemble-based inversion. Consequently, data-informed parameters are unique^40^ and quantify uncertainties^41^ associated with observation noise, waveform variability, and model resolution. Breath rate *θ* and signal baseline *y*_0_ are assumed to be stationary during parameter inference, thereby constraining the window length. Informed by these considerations, this work uses a moderate resolution model (*M* = 28) to encode and discriminate essential waveform features over 10-second windows to satisfy stationarity requirements of the inference. Each 10-second window is associated with a reference value and parameters triplet (*y*_0_, *θ*, {**a**}).

#### 2.) Parameter Distribution Summaries

Statistically summarizing waveform parameters estimates over longer stationary timescales, rather than detailing each breath, is an optional step in the pipeline. This is computationally advantageous for large dataset phenotyping because it invokes fewer pairwise comparisons of more detailed objects. The chosen parametrization process (above) samples the data-informed posterior distribution of parameters on each 10-second window. The estimators used to summarize these samples include mean, quartiles, variance, and mode, plus non-gaussian measures (skewness, kurtosis, and Kolmogorov-Smirnov distance). The latter items capture bimodal or asymmetric properties characterizing non-stationary LVS behavior. The stationary parameters, such as mean period and baseline pressure, are also included in these summary vectors. Statistical parameter summaries of 10-second windows reduce the temporal sampling rate from 31.25Hz in raw data to 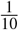 Hz while 2D raw data become ∼400-dimensional vectors of parameter estimators.

#### 3.) Including care and context information

Appending ventilator settings data to each statistical waveform parameter summary contextualizes them in the health-care process. Ventilator settings detail the mode of operation (APVCMV, PCMV, SCMV, SPONT, standby), targeted quantities (set inspiratory pressure or set tidal volume) as well as various machine settings (trigger thresholds, ramp time, mandatory minimum breath rate). Some ventilator settings such as PEEP and I:E ratio are represented implicitly in waveform descriptors and need not be explicitly included. Other available factors such as ventilator delivery power are not considered here but may be included in other applications. Ventilator mode is a nominal variable that is one-hot encoded into a set of binary variables. However, not all settings are properties of each mode. Waveform properties proxy for ventilator settings (*i*.*e*., observed maximum volume for *V*_*T*_ in PCMV), while missing data with no observable analogs (*e*.*g*., trigger settings) are filled with zero values. MV settings are included in the LVS window summaries as static properties because settings change infrequently compared with the number of windows or breaths. When available, other care-originating factors such as patient sedation level, paralytic use, and patient posture are easily included and may be used to target analysis of specific care regimes. The current implementation considers only MV settings but future effort might incorporate other care-originating factors like patient sedation level, paralytic use, and patient posture.

#### 4.) Phenotype Labeling

Phenotypes are defined through labels assigned to joint waveform-MV settings based on content similarity, which can be performed at individual patient or aggregated cohort levels. LVS descriptor vectors are reduced to lower dimensions so that labeling and assessment occurs in an easily visualized geometry. Dimensional reduction methods^42^ include analyses of factors^43,44^ as well manifold methods such as Uniform Manifold Approximation and Projection (UMAP^45,46^) and *t*-distributed Stochastic Neighborhood Embedding (tSNE^47^). Group labels are assigned by a clustering process^48,49^ applied to LVS descriptors in the reduced coordinate system that describe similarity in a non-dimensional way. Options include space partition methods (*k*-means, *k*-medoids, etc.), kernel-based methods like Support Vector Clustering^50^, and density methods like Density-based Spatial Clustering of Applications with Noise (DBSCAN^51,52^).

This study employs UMAP for reduction because it preserves local and global similarity structure of its input and has a numerically efficient MatLab implementation^53^. In this instance, the projection assesses similarity using the uniform Gower distance^54,55^ because feature vectors contain mixed-type variables. Non-uniform feature weights may be added to modulate the influence of specific data elements in future applications. For example, the impact of volume waveform components can be given less weight when investigating MV in volume control modes. DBSCAN was chosen to label groups for its ability to identify clusters of arbitrary shape, as LVS feature clouds in the dimensionless UMAP coordinates are often irregular and non-convex (Figure 1, step 4).

UMAP hyperparameters are fixed (neighborhood size 5 points, minimum distance 0.01) to maximize the equivalence of similarity-based projections across patients. During DBSCAN labeling, a brief grid-search over hyperparameters (core point requirement 4–12; neighborhood radius 1.5–5 by 0.5) finds the grouping that minimize the total distance between centroids to balance group consistency with the number identified of sets. Segmentation quality depends on data variability which increases over time; this search improves phenotype resolution uniformity across varying record lengths.

#### 5.) Phenotype interpretation

The terms ‘label’ and ‘‘phenotype’ below are synonyms because cluster labels identify consistent groups of 10-second data windows that characterize phenotypes. For example, the model images of group median parameters characterize the central behavior of pressure and volume waveforms (Figure 1, step 5 top). Every time point in an MV record carries a phenotype label, which applies to all LVS observables of the patient at that time regardless of whether they were included among features. For example, downstream analysis of *e*.*g*., FiO_2_ or SpO_2_ could use phenotype identity to stratify data.

### 2.3 Phenotypes and Characterizations of LVS data

The pipeline organizes data into discrete phenotypes based on similarity of windowed LVS states. An objective is to reveal LVS changes without corresponding ventilator settings changes. Such changes suggest the presence of factors that influence LVS trajectory including changes in patient expectation and breathing pattern (*e*.*g*., patient effort, respiratory drive), lung mechanical function (*e*.*g*., VILI progression or recovery from ARDS), or another aspect of physiology. Other factors like resistance and compliance of ventilator tubes, accumulated moisture, and changes in sedation and posture influence observed waveforms; these data are not available and remain potential confounders.

### 2.4 Experimental Phenotyping of LVS data

The pipeline described above is applied individually to 35 LVS records defining a context of 2-day periods of a few ARDS patients. This narrow context provides an opportunity to demonstrate the complexity of characterized behaviors as phenotype diversity is expected to be limited. There is no a priori reason to expect phenotypes to organize around particular data elements because the similarity metric (§2.2, item 4) weights components equally. *These experimental applications of the pipeline investigate what data elements impact phenotype structure and what variability remains in phenotypes*. The LVS trajectories are presented as a timeseries of phenotype labels contextualized by ventilator settings and shown in relation to classified PVD. Pressure-volume (pV) loop characterizations of each phenotype, computed from the median of relevant parameter estimates, provide visually summarize waveform data. Such visualizations intend to summarize key features and notable changes defining the LVS trajectory. Subsequent analysis and discussions employ principal component analysis (PCA), an empirical signal factorization based on variance minimization^43,56^. This tool reveals the degree of LVS variance occurring under during stationarity to investigate non-ventilator temporal changes not identified by segmentation.

#### 2.4.1 Cohort-scale phenotyping

Direct application of the individual pipeline to cohort data is a computationally expensive problem due to the data volume (*O*(10^6^) 10-second intervals of continuous multivariate variables). A simple alternative is to develop cohort-scale meta-labels for the population of individual phenotypes. To achieve commensurable features across the cohort, volume data (in mL) are standardized by separating the scaled magnitude (in mL/kg) from volume waveform parameters. Pressure waveforms are likewise standardized by zeroing on PEEP or other support pressure and scaling by driving (peak-minus-baseline) pressure within each window. Feature vectors for cohort clustering are individual phenotype statistics of baseline pressure, driving pressure, scaled tidal volume, estimated parameters of normalized waveform data, and associated ventilator settings. Segmentation uses UMAP-DBSCAN as in the individual case but with different hyperparameter values.

## 3 Results

The clinical data associated with ARDS patients (Table 1) are an important and practical phenotyping application because attentive MV management and patient instability instigate diverse LVS behaviors. This section reports experiment results from phenotyping individual ARDS patient data records (§3.1) and the assembly of cohort-scale phenotypes (§3.2). Within individual experiments, the temporal structure of LVS data labels is examined for consistency and resolution. Phenotypes aggregated across the cohort produce generalized LVS descriptor characterizations. PVD labels provide additional context for phenotype labels derived from ventilator settings and waveform characteristics.

### 3.1 Patient-level Phenotyping

LVS patient data are identified with 20[14] (median[IRQ]) individual phenotypes, totaling 721 groups across the cohort. Many of these patient-specific phenotypes capture less than 1% of a given patient LVS record. Over 97[3.1]% of data are captured by 8[6.5] core phenotypes that represent more than 3% of an individual record. Low-occurrence phenotypes often identify outliers and brief events such as suctioning that may be eliminated by reducing label specificity via UMAP-DBSCAN hyperparameters. Because record length varies (median[IQR] 47[37] hours), over-segmentation is needed to address the more diverse behavior of longer patient records compared with shorter ones. Over-segmentation addresses the need for record length variation (median[IQR] 47[37] hours) the more diverse behavior of longer patient records compared with shorter ones.

Phenotypes primarily organize around changes in MV settings such as tidal volume, PEEP, and ventilator mode in all experiments, but they also reflect changes in the patient-ventilator (or LVS) behavior. There is high correspondence between changes in ventilator settings and persistent changes (lasting longer than 30 seconds) in individual patient phenotype labels (SI B). Changes in settings are typically (mean(*s*2*l*) *>* 60%) reflected in label changes, with ∼ 92% of changes in PEEP, MV mode. and *V*_*T*_ inducing label changes. The former assessment is biased by few settings changes in some patients and by counting changes unlikely to discretely affect waveform behavior (*e*.*g*., trigger sensitivity or mandatory breath rates). However, label-to-settings change coherence (*l*2*s*) is much lower; less than 10% of label changes are associated with ventilator settings changes. Phenotypes capture LVS variability resulting from care-related changes to MV settings in this data-limited context.

Figures 2 and 3a–d visualize particular aspects of the low-dimensional trajectories in phenotypes for two patients (#34 and #11 of Table 1, respectively). Their cases are typical of experiments in record length ( ∼24 hours), number of ventilator settings changes, and number of identified phenotypic breaths. SI C provides additional examples that show the complexity and heterogeneity of the joint LVS-care processes in time. Each joint LVS-care record shows significant variation over time with complex and non-stationary patterns.

**Figure 2.**
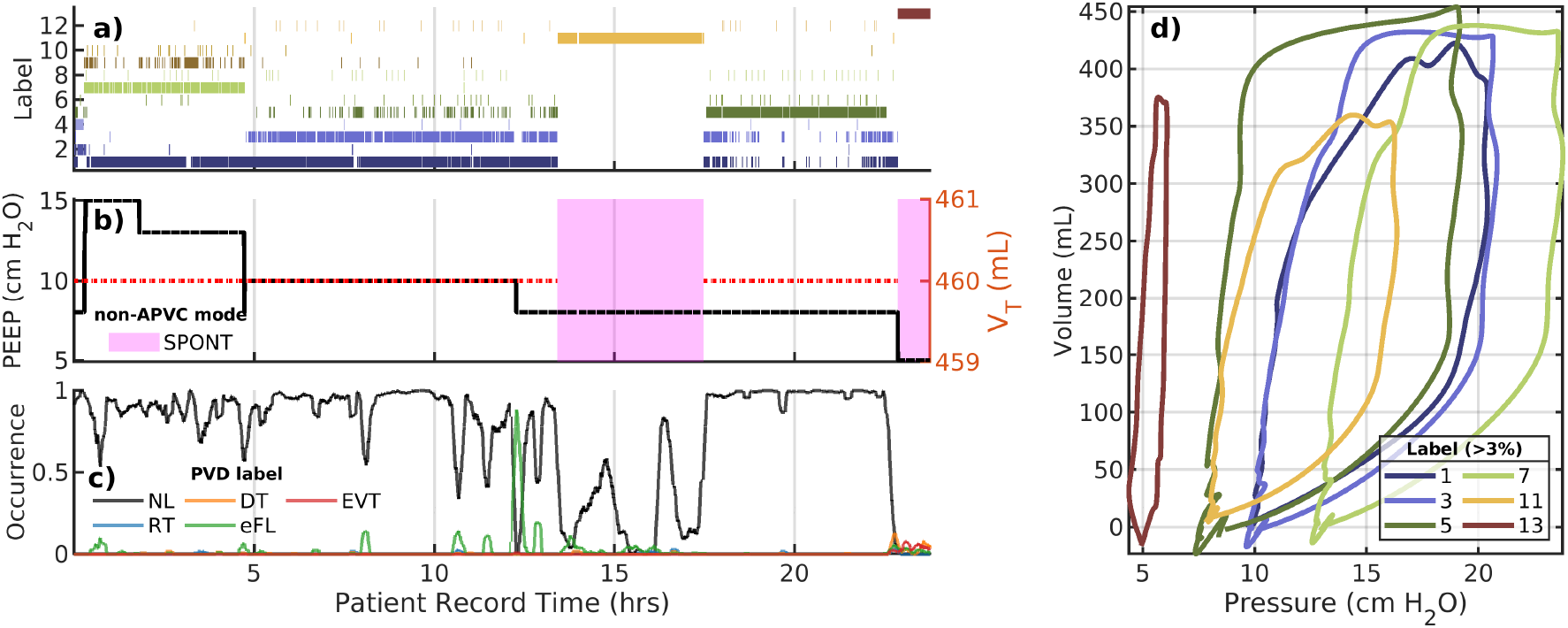
LVS evolution of patient 34. Panels a–c show the trajectory of phenotype labels, ventilator settings, and externally identified PVD, respectively with a common horizontal axis of patient record hours. In b), non-APVCMV ventilator modes are indicated by shaded regions. In c), PVD identification over time is depicted by label occurrence percentage within 1-minute moving windows. The panel (d) shows the model image of waveform parameters nearest to the group median, which characterizes breath pV loops of that phenotype (shown with the same color as (a)). The occurrence of label #1 is discontinuous in time and occurs under different PEEP values suggesting waveform shapes vary only in baseline pressure. The PVD-less evolution of the LVS shows much waveform variation separate from ventilator settings changes. Labeling and coloring in this figure do not relate to other figures.

**Figure 3.**
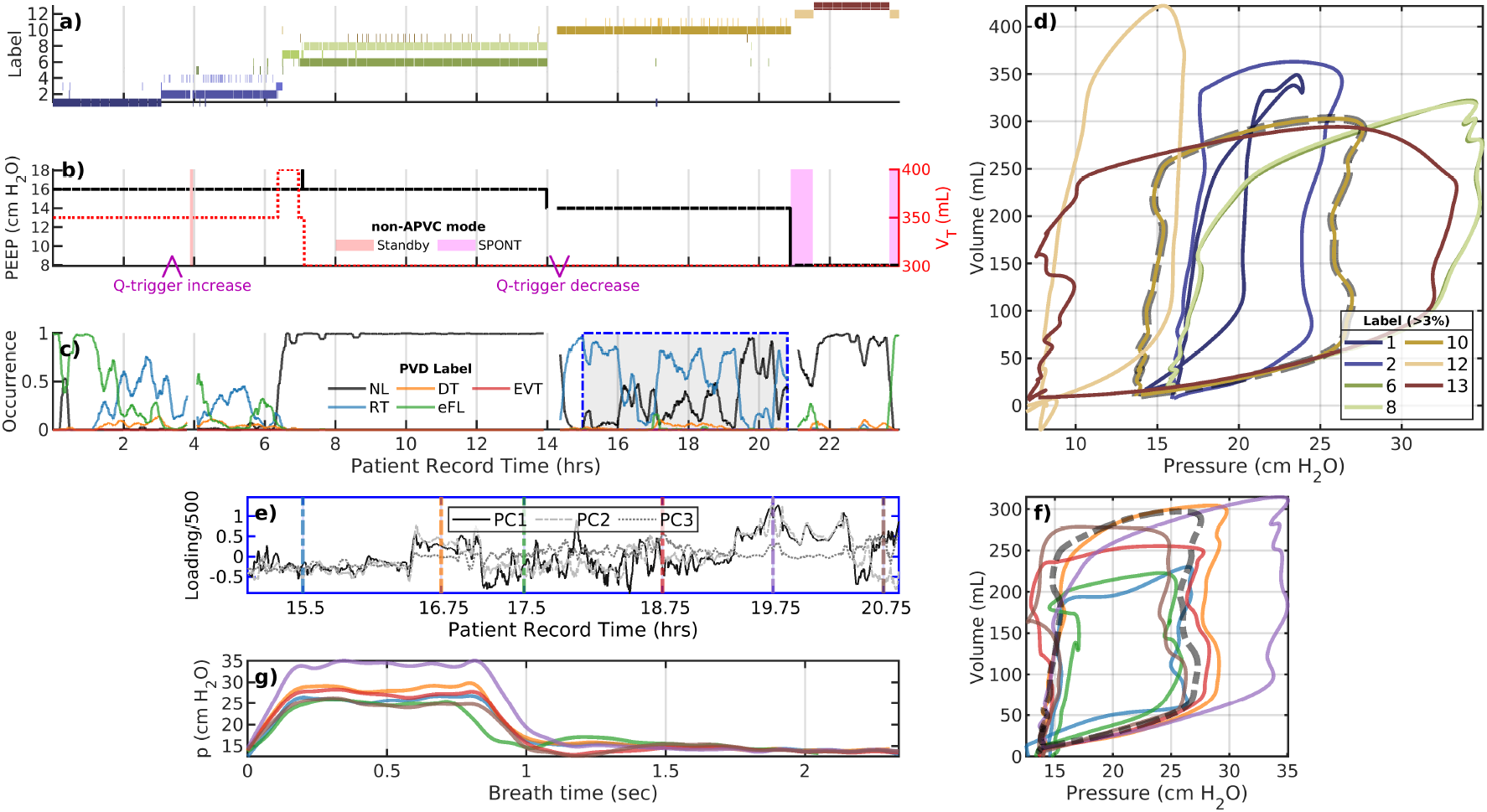
A representative example: patient 11. The plot panels (a–d) are the same as the previous figure. A flow trigger (b, at purple arrows) near 3 and 13 hours are the only MV settings changes besides mode, PEEP, or tidal volume. The lower panels (e–g) examine the variability during the record interval 15–21 hours under stationary ventilator settings. The mean (dashed black line) coincides with the golden pV loop (label #10) in the upper plot. The many distinct breath sub-types identified are more similar than to other main types in the upper plot; as a result, they are grouped together at this choice of hyper-parameters. Internal phenotype variability suggests continuous LVS changes that may not admit a natural discretization. Colors coordinate between panels a) and d), and among e), f), and g). Labeling and coloring in this figure do not relate to other figures.

#### Example 1

Figure 2 illustrates the low-dimensional trajectory of patient 35. Here, the system is driven by a progression of PEEP reductions and mode changes from APVCMV to supported spontaneous breaths for several hours. Most breaths are identified as non-dyssynchronous in externally labeled PVD (c) apart from flow-limited behavior around 12.5 hours following reduction of PEEP from 10 to 7 mm Hg. However, there is also heterogeneous behavior indicated by labels (a) during the period from 5 to 12 hours under stationary ventilator settings (b). The LVS state vacillates between labels #1 and #3 with notably distinct pV characterization (d) during this period. Irregularity in delivered tidal volume under the APVCMV and in breath length are likely explanations for variability here and in other cases.

#### Example 2

Figure 3 illustrates an analysis of patient 11 whose LVS undergoes multiple changes over a 24-hour data period. A flow trigger increase near 3 hours (b) prompts a phenotype change, and the association with eFL and RT PVD types is reflected in inspiratory coving in pV loops (d, label #1, #2, and #10). The behavior over 7–14.5 hours is identified as normal breaths characterized by quite similar phenotypes (#6 and #8); these could be merged by modifying label specificity (via hyperparameters) or via post-processing. Dyssynchronies return when the flow trigger (‘Q-trigger’) is returned to its initial value, near 14.25 hours. Breaths during brief changes to spontaneous breathing around 20 and 23 hours have markedly different pV characterizations (discontinuous label #12, tan). The interim period (20.5–22.5 hours) consists of primarily normal breaths (label #13, brown) under the default adaptive pressure volume control mode.

##### Intra-label variability: A closer look at label #10 of patient 11

The record of patient 11 (Figure 3) indicates no MV settings changes during 15–21 hours. One phenotypic breath dominates this period (c, blue dashed outline) while various PVD labels intimate variability worth scrutiny. Principal components during this interval (e) reveal structural waveform changes (f,g) that are not clearly identified as sub-phenotypes. While pressure characterizations (f) suggest the differences are attributable to pressure plateau pressure, full characterization indicates ∼ 35% variability in tidal volume (g) as well. This *continuous* variation lacks a natural discretization without altering the similarity metric, such as including other data. Figure SI 3 demonstrates a case where intra-label variability may be discretely resolved.

### 3.2 Cohort-scale phenotyping

The collection of 721 phenotypes generated from pipeline application to 35 individual records can be used to identify systemic LVS features of the cohort. Figure 4 presents key properties and labeled data distributions of 16 similarity-based clusters identified in pool of individual phenotypes. This approach to segmenting the full dataset, relying on hyperparameters and not generalizing from these data, further minimizes variability while displaying consistent waveform properties and ventilator settings. Labels mix patients (b) while separating PEEP (c), with exceptions for uncommon ventilation modes (d) applied to few patients. Table 2 and Figure 4 quantitatively validate labeling of original data in the general settings. Specifically, labels consistently align with structured properties of the LVS data. Figure 5 shows the associated non-dimensional waveform characterizations; PEEP, tidal volume, and peak pressure features are used to normalize these waveform data elements across patients.

**Table 2.** Cohort label properties. Columns identify: cohort-level label, percentage of 10-second windows, number of contained patients (*N*_pat_), number of contained individual phenotypes (*N*_pheno_), median[IQR] of baseline pressures (*p*_base_, typically PEEP) and pressure change (Δ*p* := *p*_peak_ − *p*_base_) in cm H_2_O, median[IQR] of tidal volumes (*V*_*T*_ ) in mL/kg, and dominant associated ventilator mode. Values are determined from breath-level source data whose individual phenotypes share a given cohort phenotype. *=5–10% SPONT, **=10–20% SPONT, ***= 40% SPONT

| Label # | Total% | $N_{\text{pat}}$ | $N_{\text{pheno}}$ | $p_{\text{base}}$ | $\Delta p$ | $V_T$ | $\Delta p/V_T$ | MV mode |
| --- | --- | --- | --- | --- | --- | --- | --- | --- |
| 1 | 15.5 | 23 | 101 | 10 | 12.1[3.7] | 6.3[1.0] | 1.9[0.6] | APVCMV |
| 2 | 13.8 | 22 | 101 | 12 | 14.2[3.3] | 6.0[1.1] | 2.3[1.0] | APVCMV |
| 3 | 11.4 | 11 | 52 | 8 | 12.7[4.1] | 7.9[1.3] | 1.5[0.4] | PCMV* |
| 4 | 8.4 | 12 | 37 | 14 | 12.2[6.9] | 5.9[0.2] | 2.0[1.3] | APVCMV* |
| 5 | 7.3 | 11 | 32 | 12 | 15.1[13.6] | 5.9[0.1] | 2.7[2.4] | APVCMV |
| 6 | 6.9 | 17 | 58 | 11 | 13.1[2.9] | 6.2[1.3] | 2.1[0.5] | APVCMV |
| 7 | 6.3 | 8 | 49 | 14 | 12.6[2.9] | 6.2[1.3] | 1.9[0.7] | APVCMV |
| 8 | 6.2 | 11 | 49 | 16 | 13.4[2.3] | 6.0[0.6] | 2.2[0.6] | APVCMV |
| 9 | 6.2 | 9 | 51 | 16 | 15.9[6.0] | 5.9[2.8] | 2.6[2.4] | APVCMV |
| 10 | 4.1 | 11 | 34 | 8 | 9.7[2.7] | 6.8[1.5] | 1.6[0.4] | APVCMV |
| 11 | 3.7 | 5 | 25 | 5 | 10.7[0.2] | 6.5[0.7] | 1.7[0.2] | PCMV** |
| 12 | 3.4 | 14 | 22 | 5 | 8.9[4.1] | 7.0[2.4] | 1.2[0.8] | APVCMV*** |
| 13 | 2.5 | 6 | 10 | 8 | 11.1[1.4] | 6.6[1.0] | 1.7[0.2] | APVCMV |
| 14 | 1.7 | 11 | 27 | 14 | 13.3[2.9] | 6.0[1.3] | 2.0[0.8] | APVCMV |
| 15 | 1.5 | 5 | 14 | 10 | 13.5[1.9] | 6.5[0.3] | 2.1[0.4] | APVCMV |
| 16 | 1.1 | 10 | 16 | 14 | 21.3[9.5] | 5.6[1.8] | 3.7[3.1] | APVCMV |

**Figure 4.**
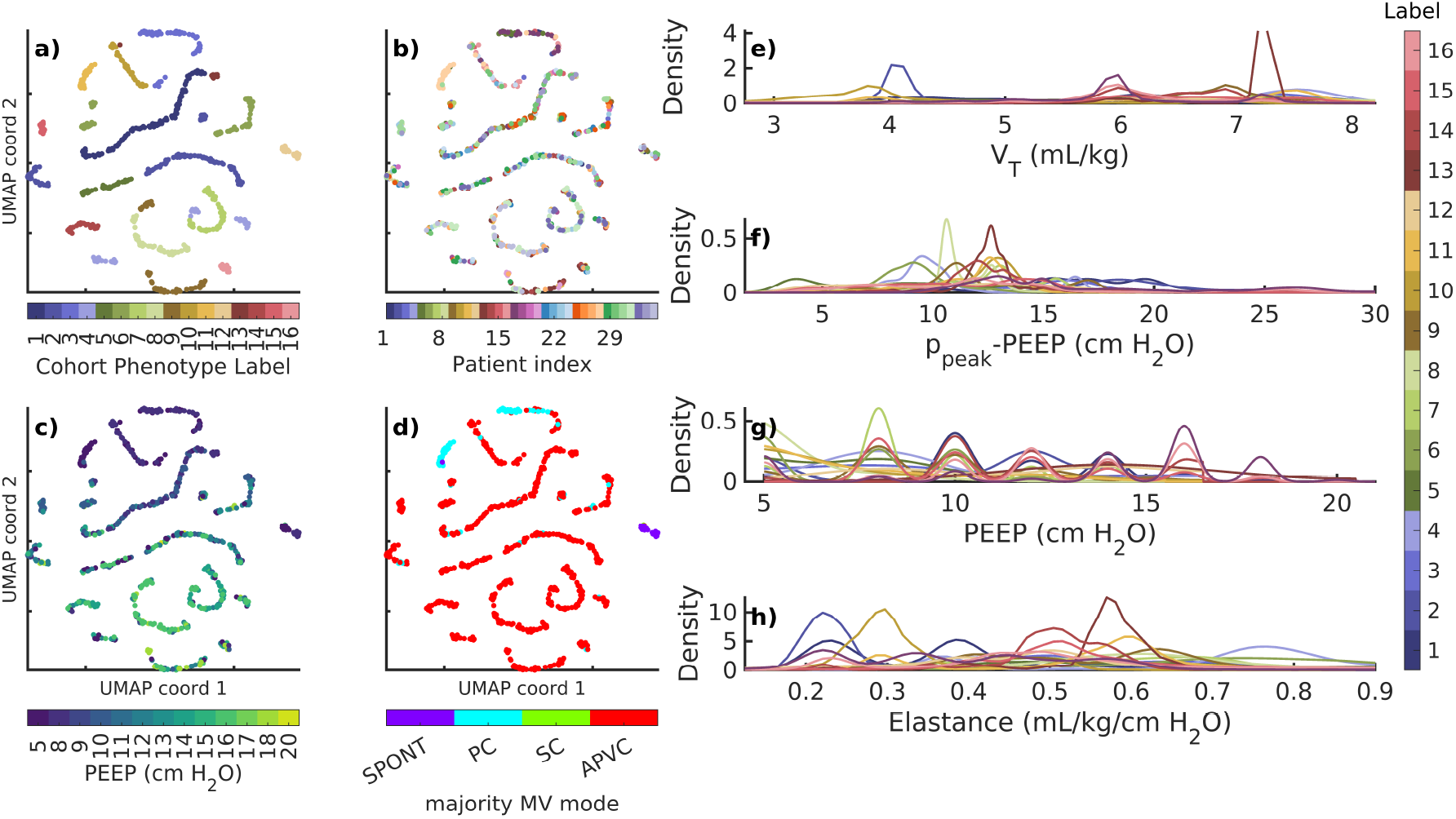
Membership and data properties associated with cohort phenotypes. Points in panels (a–d) correspond to 721 individual phenotypes shown in unitless 3D UMAP coordinates that describe similarity (only two axes depicted, for simplicity). Labels (a) mix patients (b) while defining empirical partitions of other factors of patient data (c–h). Groupings separate PEEP (c,g) and ventilator modes (d), which are arguably among the most important ventilator feature elements. Structured distributional separation occurs for continuous breath variables such as tidal volume (e), driving pressure (f), and elastance (*V*_*T*_ */*(*p*_max_ − *p*_base_), g). PEEP (c) and ventilator mode (d) of UMAP labels identify the median value of each individual phenotype; probability densities (e–h) are computed from original data and colored according to panel (a). Labels and colors of panel a) define the those of panels e–h) and Figure 5. *Modes: spontaneous (SPONT), Pressure controlled (PC), Synchronized controlled (SC), and Adaptive Pressure Volume Controlled (APVC)*

**Figure 5.**
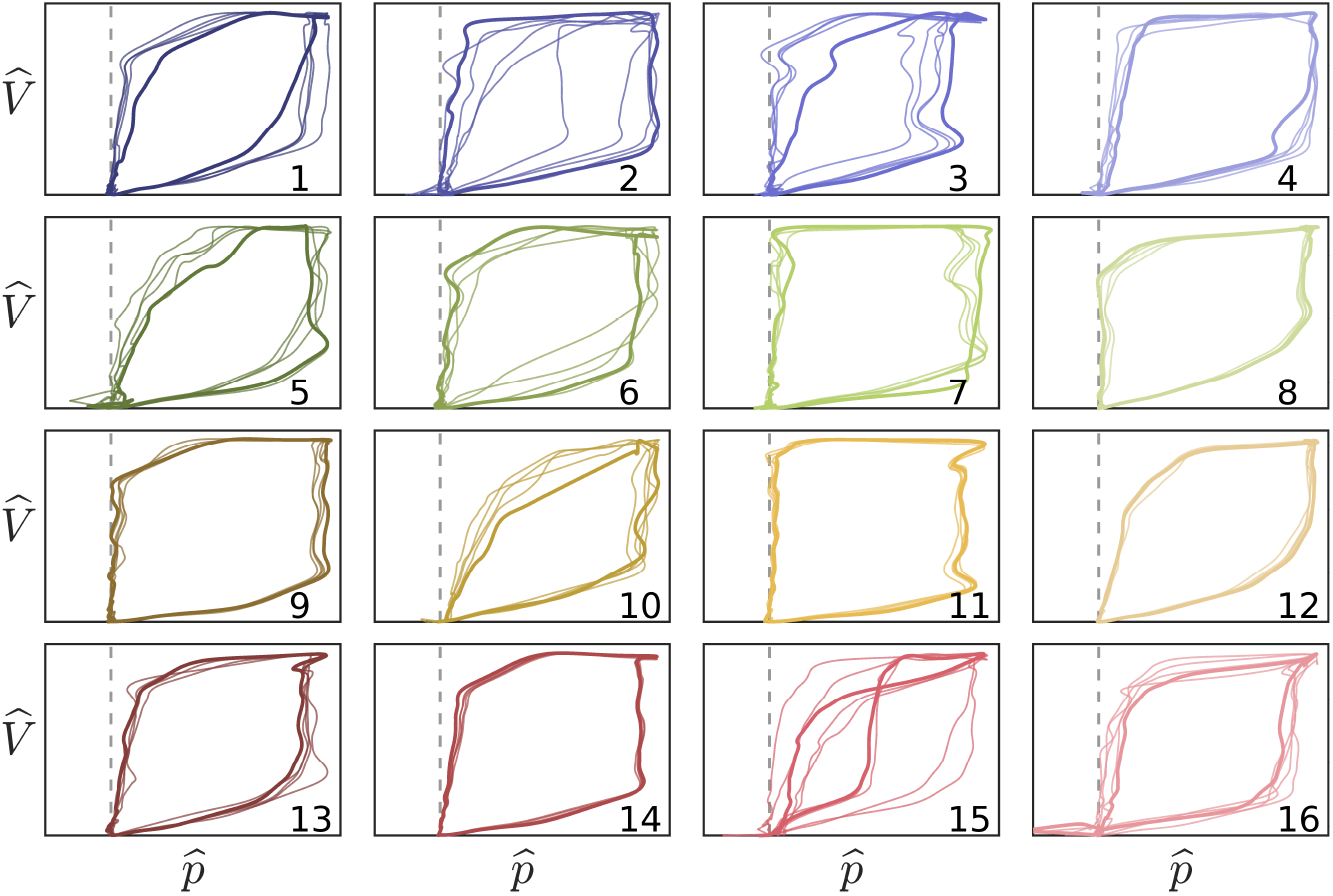
Non-dimensional waveform characterizations. Pressure-volume traces correspond to median (bold) and nearby (thin) window characterizations of each cohort phenotype. Labels and colors correspond to Figure 4a. Vertical and horizontal scales axes correspond to 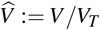 and 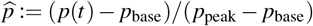, respectively, per Figure 4e–g. The dashed line indicates baseline pressure. Cohort phenotypes differentiate waveform shape characteristics and pressure-volume coordination in conjunction with associated scaling factors. Intra-group variation is naturally high given the low specificity of each type.

Granularity of cohort meta-characterization depends on UMAP-DBSCAN hyper-parameters (UMAP: neighborhood size 12, minimum distance 1; DBSCAN: epsilon 2.7, min points 5). The small sample size (*N* = 721) lead to robust UMAP representation but high sensitivity to neighborhood size in DBSCAN (SI D). Chosen parameters aimed to maximize the number of phenotypes while easily communicating waveform characterizations in an array of figures; the results are qualitatively similar for nearby parameters. Table 2 summarizes the occurrence and properties of the 16 cohort phenotypes.

### 3.3 Synthesis

Experiment results provide insight into the structure and heterogeneity of empirically phenotypes constructed for 2-day ARDS patient LVS+care data. Based on equally weighted data elements, these groupings primarily organized around ventilation mode and PEEP followed by tidal volume and internal LVS variability, giving a hierarchical organized by key MV settings and waveform instability. LVS behavior is shown to vary in several ways under MV settings stationarity that may be of particular interest for VILI detection and tracking of ARDS progression. This local variability can be resolved by phenotypes as in Figure 2a, during hours 5–12, 17–18, 22–23) and Figure 3a (hours 3–14). Groupings can also mask important breath heterogeneity if phenotypes are too coarse (*e*.*g*., Figure SI 1e–g and Figure SI 3. Resolution of behavior categories is improvable through UMAP-DBSCAN parameter optimization or local analysis, such as application of PCA. Heterogeneous phenotypes (*e*.*g*., Figure 3e–g) also arise in diverse groups of breaths with continuous local similarity (*e*.*g*., Figure 1, step 4, label #4 in red). Weighting feature components in the similarity metric can improve sub-type resolution of such behaviors. Additionally, cohort scale analysis is easily computed from a cohort of individual phenotypes. The resulting phenotypes are specific to the data and algorithm parameters and do not generalize, but the modular pipeline process may be adapted and manipulated for more specific purposes. Such categorization provides a coarse but scalable and unified basis for analyzing the evolution of LVSs in terms of their consistent statistical properties.

## 4 Discussion

This study presents a framework for extracting meaningful, low-dimensional characterizations of lung-ventilator system (LVS) states from observable data of managed patient-ventilator systems. Consequently, the observable LVS data, whose contents define a limited context of application, are reduced to a discrete set of patient-level phenotypes. The phenotyping pipeline was developed as a generalized process to accommodate many methodological choices and hypotheses about which LVS factors are important in a given application. This general methodology contrasts with the construction of a generalized output, as source data insufficiently sample the breadth of MV processes including other ventilators, sites, patient characteristics, and care protocols. The approach instead provides a practical means for researchers to explore LVS evolution from data with significant heterogeneity caused by ventilator adjustments and changes in patient-ventilator dynamics. In particular, the data-specific phenotypes naturally disentangle effects of MV settings from the human-machine interaction of the LVS. Research toward improving and personalizing MV benefits from phenotypes that include both ventilator settings and patient-ventilator interaction. Both LVS factors are necessary to accurately analyze MV consequences in light of patient and time heterogeneity. The phenotype identification process is intended for research rather than for MV decision support. The need for a framework to develop hypotheses about temporal effects of MV and local outcome validation targets from retrospective data motivated the developments presented in this work.

Experiments on 35 ICU ARDS patients, including 8 COVID-19 cases from 2020, performed segmentation with uniform LVS feature weights. Fixed similarity assumptions and hyperparameter optimization ranges were used to define individual-scale phenotypes, which were later compared in cohort-scale phenotyping. LVS categories reflected null hypotheses that prioritized no particular features of the included data in phenotype definition. Individual results showed that phenotypes primarily organize by ventilator mode, PEEP, and tidal volume. This effectively separated care processes from the patient-ventilator component of the data generating system, while analysis of periods under persistent VM settings showed LVS changes to be more complicated. Whereas MV settings changes are abrupt, LVSs exhibit a variety of behaviors including continuous but non-monotonic progression (Figure 3), transient behavior (Figure SI 1), and alternation between both similar and non-similar breath patterns (Figure 3). Phenotype resolution could be adjusted to further delineate certain discrete variations, while continuous changes resist discretization without feature weighting or other source data. Continuous changes may result from apparatus properties (such as changes in tube compliance, resistance from accumulated moisture and bends, and leakage) as well as effects of patient sedation. However, they may also suggest progressive effects of lung physiology under MV. Investigating such behaviors first requires identifying categories, like those developed in this work, for which heterogeneity can be calculated.

### 4.1 Validation and Interpretation

The created typologies are based on similarities among observable data, making them phenotypes of patient-ventilator-care data representative of LVS behaviors. Clinical validation of output requires quantitative comparison with patient state or conditions^57^, but such biomarkers of breath behavior do not currently exist. Further, global outcomes (discharge disposition, 30-day mortality, *etc*.) are unlikely to relate to local behaviors observed during 1–2-day segments of MV encounters. Investigation demonstrated label consistency in relation to changes in PEEP, ventilator mode, and tidal volume for phenotypes based on naive hyperparameters and an uninformative similarity metric. The analysis qualitatively validates practical application by identifying what LVS behaviors computed phenotypes did and did not differentiate as well as what variability can be isolated via hyperparameter tuning.

Phenotypes are more granular than a ventilator settings-based classification (Table SI 1) and more generalized than PVD labels, which target specific behaviors. Label heterogeneity suggests potential label sub-types so that hierarchical or multi-stage clustering are important refinements in future applications. Although 10-second window scalar phenotypes are directly incomparable to breath-wise vector types of PVD, changes in label-described behavior strongly coordinate with changes in PVD type. Notably, phenotype variability analysis and PVD labels identified qualitatively similar temporal patterns (*e*.*g*. §3.1 and SI C) without dyssynchrony labels informing LVS descriptors. Additionally, esophageal pressures were not encoded into phenotypes but are required to confirm certain PVD types^21^. Cohort labels demonstrably partition data into groups albeit with an expected high degree of variability given the reduction of ∼1.5M breaths to 16 categories. Their identification required waveform component normalization to ensure patient comparability that would benefit from stratified analysis based on mode, PEEP, and primary control variables. Despite the coarseness of categories, signs of dyssynchrony are apparent in these median pV shapes such as ineffective triggering (sub-baseline pressures in #5, #15, and #16) and flow limitation (inspiratory coving in #3, #11, and #15). This indicates that some of the cohort scale phenotypes, while broader and less specific than PVD types, center on elements of dyssynchronous behavior. Including PVD labels or other physiological information in feature descriptors may better align phenotypes with PVD labels in applications targeting LVS specific behaviors.

### 4.2 Innovations, Limitations, and Improvements

This work discretized joint patient-ventilator-care system data as holistic units to overcome limitations on analysis imposed by data complexity and heterogeneity. This phenotyping approach is generalizable, suitable for other datasets, and can accommodate different feature and clustering options. The process and results are geared toward data-driven research use rather than clinical informatics or clinical decision support. In scientific application, practitioner guidance is required to inform data features and their importance relative to target observable investigated via LVS phenotypes. Nevertheless, the developed method and specific implementation assumes certain conditions and has limitations.

The presented phenotyping pipeline outputs are neither generalizable nor clinically validated. This is a consequence of insufficient data to sample all LVS behavior, lack of a “gold standard” MV breath typology, and absence of LVS state biomarkers. Target biomarkers would aid in feature design of the pipeline through clinical knowledge and physiology regarding how such observables relate to LVS data used here. What the phenotyping process provides, however, are categories to which clinical consequences may be attributed in further study.

The pipeline ignored uncommon esophageal pressure data, which are essential to confirm certain dyssynchronies, because they require high model resolution to resolve and have inconsistencies (gaps, drift) that limit continuous time characterization. However, these data were used to manually identify PVD in breaths used in the supervised PVD labeling^22^ featured in validation. The waveform parametrization also relies on ventilator-identified breath cycles, so the pipeline lacks the flexibility needed to identify double-triggered PVD events that occur over multiple ventilator cycles. Analysis omitted important potential influences such as neuromuscular blockade use, position/posture, and airway secretions whose data were not available. LVS descriptors easily can incorporate these factors to better resolve care-stationary periods and more precisely resolve LVS variability. Additionally, data reflect only one ventilator model; additional harmonization is needed to compare breaths generated by different ventilators because mode settings and pV observation points may differ.

Finally, the group identities of empirical phenotypes depend directly on hyperparameters that govern similarity and specificity. Fixed UMAP parameters reflected a constant local similarity assumption, while DBSCAN parameters were optimized over a narrow domain to account for differences in record length. The dimensional reduction process employed a similarity metric with uniform feature weights to limit external assumptions. Practical applications should incorporate background hypotheses to target feature weights that emphasize key LVS data features of interest. Further improvements can easily involve outer-loop targeting of application-specific objectives beyond the generalized scope of this work.

### 4.3 Concluding Remarks

This work developed a flexible categorization process for context-constraining data timeseries from patient-ventilator system under managed care. The research outlined a process of empirically discretizing relevant observational data capable of isolating patient-ventilator dynamics from care processes and labeling data subsets based on similarity. Assessing phenotype local heterogeneity is an essential first step in temporal analysis of MV patient data within the context of applied care. Ongoing work toward formulating hypotheses about system trajectories related to applied care, local variability, and outcome motivated providing a shared low-dimensional basis for LVS comparison, which also motivated the construction of cohort-scale phenotypes. These ongoing efforts are inextricably linked with clinical validation, which requires that quantified clinical consequences of MV be tied to phenotypes through hypotheses involving both phenotype definitions (*viz*., the data and pipeline choices defining them) and the nature of behavior-to-consequence association. Converting LVS dynamics to sequential progression through finite states allows the application of symbolic dynamics^58–60^, game theory^61^, discrete-time Markov chains, and large language models. Such tools can extend this works’ investigation to patient trajectory patterns and their consequences improve understanding of dynamical effects of current MV protocols on patient-ventilator systems.

## Acknowledgments

This work is supported by National Heart Lung and Blood Institute awards 5R01HL151630 “Predicting and Preventing Ventilator-Induced Lung Injury” (DJA), K23HL145011 “The Detection, Quantification, and Management of Ventilator Dyssynchrony” (PDS), and K24HL168225 “Mentoring and Patient-Oriented Research in Clinical Informatics and Data Science” (TDB). Thanks as always to Meg Rebull for local administrative support.

## Declaration of Generative AI and AI-assisted Technology Use

ChatGPT 3.5 provided suggestions for rephrasing complex sentences in early drafts. No other AI-assisted technologies were employed and no generative model output was used directly.

## Author Contributions

DJA, JNS, YW - methodology and conceptualization; PDS, JNS - formal analysis; JNS - development, writing, and visualization; DJA, MM, BJS, PDS - project management; MM, PDS - data acquisition and preparation; DJA,TDB, BJS - resources and funding. All authors reviewed the manuscript.

## Data Availability

The clinical datasets used and/or analyzed during the current study are not publicly available due to ongoing collection, lack of patient consent for broad dissemination of their data, and data size (25 GB). Data are available by reasonable request to the author (J.N. Stroh) and will require a data use agreement with the data owners.

## Supplemental Information

**ABSTRACT**

A set of supplemental details to accompany the main text.

## SI A Inference of model-based parameters

The waveform parametrization adopted in this work is briefly outlined below; further details, error analysis, and validation of the parametrization are found in prior publication^1^. Briefly, the model transforms waveform data *y*^obs^ on a continuous time window *I* into a parameter vector **a** under the assumption that PEEP and breath period *θ* are locally constant. A differential equation models the state variable *y* (pressure, volume, or another waveform variable of interest):

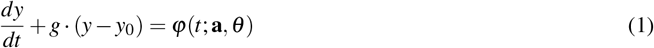

where *t* is time, *g* is a smoothing parameter, *y*_0_ is baseline or reference state (such as PEEP when *y* is pressure, and zero when *y* is volume), and *ϕ*(*t*) is a time-dependent function of local amplitude parameters *a* ∈ ℝ ^*M*^ and breath period *θ* . The model, Eq(1), defines a map **a ↦** *y*(*t*) that simulates a state trajectory *y* from a periodic step function *ϕ* modulated by parameters **a**:

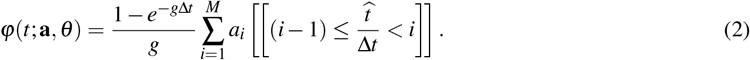

where 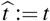 (mod *θ* ) is time within the breath, which is divided into *M* equal epochs of width Δ*t* = *θ/M*. The function *ϕ* is piecewise constant in Eq2. That is: on interval 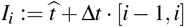, Eqn (1) is simply *y*^′^ + *gy* = *C*_*i*_ for some constant *C*_*i*_ (regardless of whether epochs are equal).

Structurally,error the model can be written as *y*^′^ + *gy* = *f* (*t*; *ω*) where *ω* = (*a, y*_0_, *θ* ) ∈ **R**^*M*+2^ is the full parameter vector. The function *f* (*a, y*_0_) = *ϕ*(*a*) − *gy*_0_ is a simple linear function that is constant on subintervals {*I*_*i*_ : *i* = 1..*M*} that partition the local breath domain *I*. Thus on each subinterval, the equation is *Ly*_*i*_ = *C*_*i*_; *y*(0) = *y*_*i*,0_ where 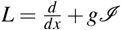 is the linear differential operator, *C*_*i*_ is the value of *ϕ* on *I*_*i*_, *y*_*i*,0_ is the initial data for *I*_*i*_, and ℐ is the identity function.

The initial value problem has a unique solution

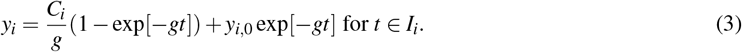

The solution on *I* is found by solving over *I*_*i*_ in sequence using terminal values as initial data for the next subinterval. That is, *y*_*i*+1,0_ = *y*_*i*_(*T*_*i*_) where *T*_*i*_ is the left endpoint of *I*_*i*_ for *i* = 1..*M* − 1, and *y*_1_(0) := *y*^*obs*^(0) This solution *L*^−1^ *f* is absolutely continuous and uniquely specified by parameters.

### SI A.1 Parameter identification via model inversion

The model defines a map from parameters to trajectories, but what is desired to parameterize data is a map from observable data to parameter distributions *y*^*obs*^ ↦ {**a**} which requires model inversion^2^. Optimal parameter distributions for pressure and volume data are generated by applying an ensemble Kalman-like smoother using a moderate resolution model (*M* = 28) to sequentially estimate parameters for each 10-second block of data using 1.6 second windows with 0.8 second overlaps.

Specifically, a windowed application of the Ensemble Kalman Smoother (EnKS)^3^ is used for parameter estimation. This technique approximates a Bayesian update by minimizing the discrepancy between a model forecast trajectory and corresponding external observations 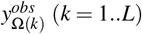 for the *k*th window Ω(*k*) using an ensemble forecast over the local times {*t*_*k*_}, *k* = 1..*L*. The target parameters **a** are those that minimize the standard 4D-var cost function^4^

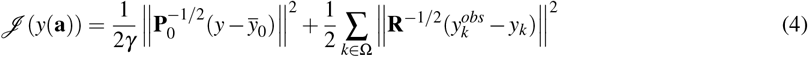

Here, *k* indexes time points of data within an inversion subwindow and *γ* is a covariance inflation factor that adjusts the weight of the data in the analysis. The initial data and parameters for the ensemble are characterized by the mean 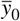 and matrices 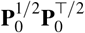, and **R** = **R**^1*/*2^**R**^⊤*/*2^ represents model and observation error covariances, respectively . The symbols 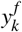 denote simulated observations generated by the model using the initial state *y* and parameters **a**, and Ω is a subset of time indices for data assimilation. The ensemble of parameter variations around the optimal mean is retained to reinitialize the model forecast at time *t*_*S*_ (0 *< S* ≤ *L*) for the next forecast-correction iteration.

Computationally, the ensemble state anomaly matrix **P**^1*/*2^ is constructed column-wise by deviations from their mean and scaled by 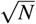. A feature of ensemble-Kalman type approximations to Eq(4) is that each potential solution lies within the subspace 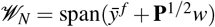. Therefore, each possibility is determined by a control vector *𝓌* ∈ R*N* where *N* is the number of elements in the ensemble. Exploiting this detail, an optimal subspace solution 𝓌 ^∗^ is defined as the variable that minimizes the cost function over *𝒲*_*N*_ . This is given by

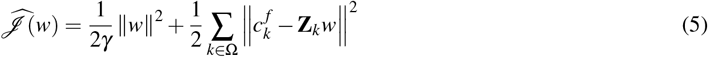

where 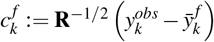 are the mean forecast error and **Z**_*k*_ := **R**^−1*/*2^**P**^1*/*2^ are ensemble forecast anomalies at the time of observation *y*_*k*_. This has a simple quadratic form

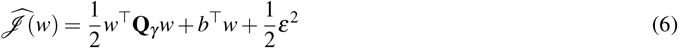

where 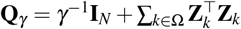 is the Hessian matrix, *b* := −∑_*k*∈Ω_ **Z**_*k*_*c* ^*f*^ is the linear coefficient vector, and 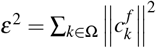 is the sum of squared differences between the initial ensemble forecast and data. Here, **I** is the identity matrix and (·)^⊤^ denotes the matrix transpose. The minimum of the cost function is characterized the vanishing gradient of 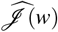 with respect to *w*. The corresponding solution of 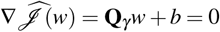 is the optimal coefficient

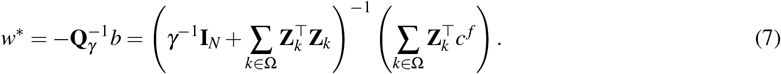

**Table SI.1.** *N*_*s*_ indicate the number of ventilator settings changes in set_PEEP, set_ptrigger, set_qtrigger, set_rate, set_fio2, set_ie, set_flowpat, set_mode, set_vt, and vt_set. *N*_*l*_ indicates the number of persistent label changes, counting those lasting longer than 30 seconds, to omit isolated transient changes and variability occurring as mixed-breath types (*e*.*g*., Figure SI 2 during 11–14 hours, characterized by both alternation between labels #10 and #13 labels and changes in ML-identified PVD type). Column ‘s2l’ indicates the percentage of vent settings changes that occur with a label change within 100 seconds. Column ‘l2s’ indicate the number of label changes that occur within 100 seconds of vent changes.

| Patient | $N_s$ | s2l (%) | $N_l$ | l2s (%) | ID | $N_s$ | s2l (%) | $N_l$ | l2s (%) |
| --- | --- | --- | --- | --- | --- | --- | --- | --- | --- |
| 1 | 45 | 73.33 | 66 | 16.67 | 18 | 11 | 45.45 | 138 | 3.62 |
| - | 1 | - | 1 | - | 19 | 14 | 85.71 | 296 | 6.42 |
| 2 | 2 | 100.00 | 35 | 8.57 | 20 | 10 | 70.00 | 25 | 20.00 |
| 3 | 25 | 36.00 | 356 | 1.97 | 21 | 37 | 78.38 | 154 | 18.83 |
| 4 | 1 | 100.00 | 46 | 4.35 | 22 | 76 | 5.26 | 175 | 0.57 |
| 5 | 99 | 56.57 | 288 | 9.72 | 23 | 104 | 86.54 | 695 | 6.47 |
| 6 | 20 | 75.00 | 342 | 2.92 | 24 | 222 | 59.01 | 590 | 6.78 |
| 7 | 3 | 33.33 | 73 | 1.37 | 25 | 137 | 54.01 | 166 | 14.46 |
| 8 | 23 | 78.26 | 328 | 7.01 | 26 | 10 | 60.00 | 38 | 21.05 |
| 9 | 59 | 45.76 | 177 | 6.21 | 27 | 24 | 70.83 | 451 | 3.99 |
| 10 | 14 | 78.57 | 260 | 6.15 | 28 | 47 | 100.00 | 629 | 1.75 |
| 11 | 48 | 29.17 | 50 | 24.00 | 29 | 73 | 45.21 | 431 | 7.42 |
| 12 | 0 | - | 4 | 0 | 30 | 37 | 62.16 | 713 | 2.66 |
| 13 | 83 | 80.72 | 370 | 12.70 | 31 | 83 | 77.11 | 421 | 9.03 |
| 14 | 13 | 76.92 | 1000 | 1.40 | 32 | 50 | 62.00 | 380 | 8.42 |
| 15 | 51 | 98.04 | 265 | 8.30 | 33 | 220 | 42.27 | 650 | 9.38 |
| 16 | 57 | 40.35 | 246 | 8.94 | 34 | 39 | 25.64 | 296 | 4.73 |
| 1 17 | 18 | 88.89 | 460 | 3.91 | 35 | 12 | 75.00 | 424 | 2.12 |
|  |  |  |  |  | mean | 52 | 64.57 | 325 | 8.00 |

Once the optimal trajectory *y*^∗^ = *y* ^*f*^ + **P**^1*/*2^*w*^∗^ is known, so are the model parameters used to generate it. To update the prior distribution for the next forecast window, the ensemble variations are recentered around this optimal solution point at some future time point *t*_*S*_ by reweighting the forecast covariance factors by those of the inverse Hessian matrix: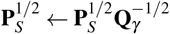.

## SI B Label changes in relation to vent settings changes

Table SI 1 shows that most vent settings changes are accompanied by changes in labels. However, very few phenotype label changes correspond to changes in vent settings. *Over 64% of ventilator settings changes are identified in label changes*. A larger proportion (*>*87%) are identified when limited to PEEP, tidal volume, and model changes, which induce significant waveforms changes compared with other settings such as mandatory breath rate (set_rate). *Few (8%) identified changes in label, however, are directly associated in time with ventilator settings changes*.

## SI C Individual Experiments, continued

This supplement continues illustrated examples of §3.1.

Figure SI 1 panels a–d illustrate the analysis of Patient 2 whose data consists of 7 record hours with one simple ventilator setting change. Only ventilator PEEP (a) is changed while there are three primary behaviors identified (b,d). The reduction of PEEP occurs about 2 hours following a rise in early flow limited breaths (eFL, panel c). This PEEP change (from 8 to 5 cm H_2_O) shifts peak pressure from 16 to 12 cm H_2_O for about an hour, at which time higher esophageal pressures returns. These breaths are identified as normal (NL)^5^. Increased specificity may be pursued by local segmentation or other dimensional reduction methods.

### A closer look at label 1 of patient 2

The first principal component loadings (panel e, black) for LVS descriptors over the first 5-hour period track the sequence of normal and eFL PVD labels (f, shown as 5-minute statistics for clarity). Direct correlation between continuous loading values on 10 second windows and statistical breath-wise binary PVD label is not well-defined while binary-to-binary comparison is. Here, the sign of loading for the first principal component is compared with PVD label. Within the same breath phenotype (label 1), the *sign* of the component loading statistically the eFL PVD labels (AUROC=0.8718); high positive values are associated with eFL breaths (f,g; green) where pressure maxima proceed volume maxima. These LVS variations result from changes in the patient component, as there is no change of ventilator settings.

**Figure SI1.**
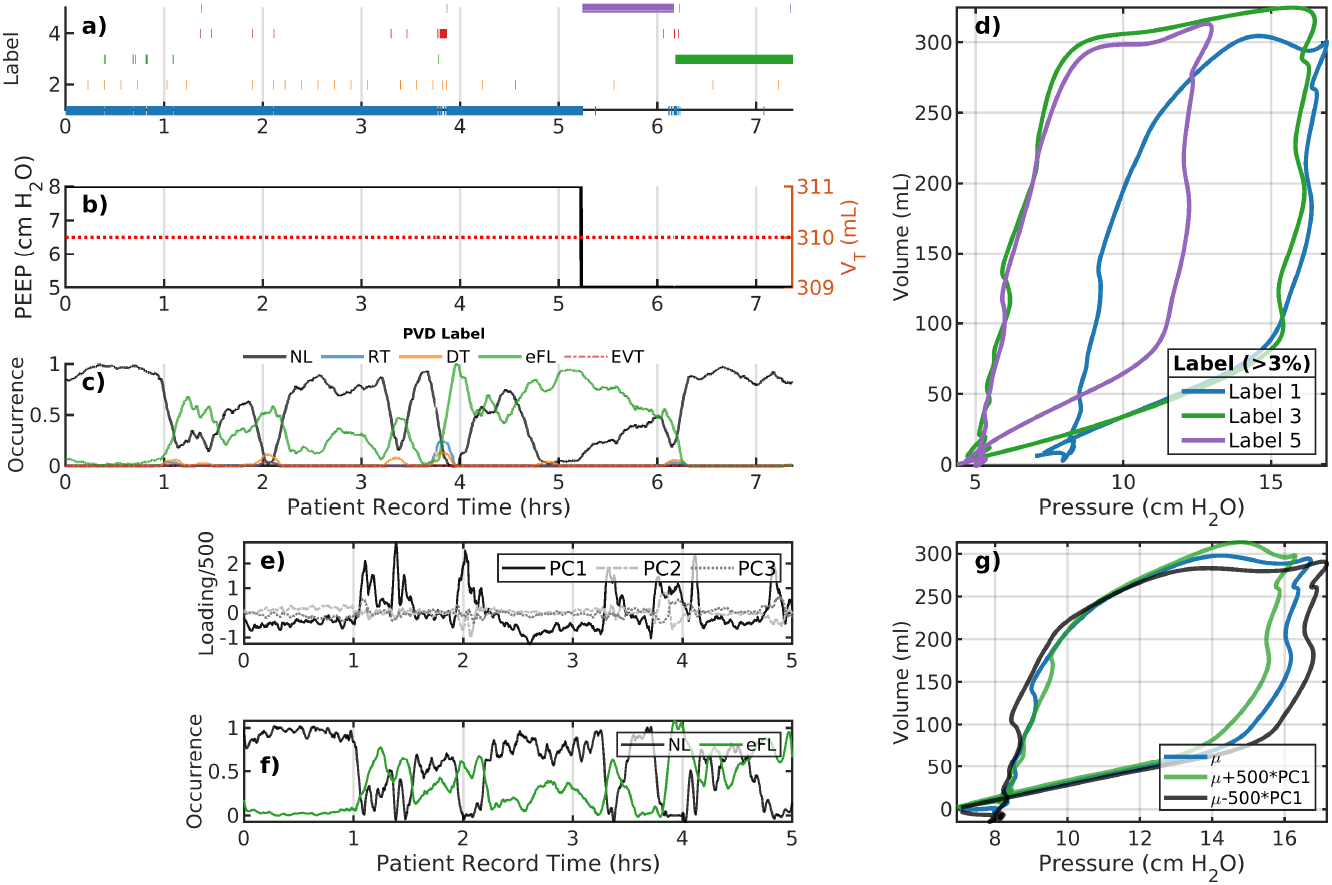
Analysis of patient 2 LVS data (a–d) and the initial a 5-hour interval (e–g). Panels a–c correspond to changes in ventilator settings, segmentation labels, and identified PVD type, respectively. The horizontal axis for these panels is the patient record time in hours. The panel (d) shows the model image of segmented data median parameters, which characterize the pV loops of breaths with that label (shown with the same color). Evolution of the LVS can be parsed pictorially from these figures. Large positive variations in the first principal component loading (e, black) for the initial 5-hour period align with PVD labels indicating eFL type breaths (f) for this period. Specifically, this suggests discrimination of breaths shapes (g) can be differentiated using qualitatively criterion on local loadings or other segmentation. Labeling and coloring in this figure do not relate to other figures.

The patient 10 (Figure SI 2) dataset is nearly twice as long with again only one PEEP change occurring after 10.5 hours of the 15.6 hour record. Breaths are stably identified as normal-type until about 8 hours, occupying two cluster-identified similar breath shapes. This is followed briefly by eFL breaths and a transition to a new characterization (label #8, light green) for about 30 minutes. In the following period (9–14 hours), breaths are characterized by lower pressure maxima (label #10, gold); these are associated/identified with reverse-trigger breaths (primarily RTm) and waveforms featuring pronounced inspiratory pressure drop. The reduction in PEEP slightly increases the incidence of normal breaths during 11–14 hours although this results in the more frequent appearance of shallow breaths (label #13, red).

#### SI C.1 Intracluster variability in relation to PVD labels

This final example shows a case where principal component loadings strongly tie to difference between normal and eFL breaths in patient 8, label #2.

**Figure SI2.**
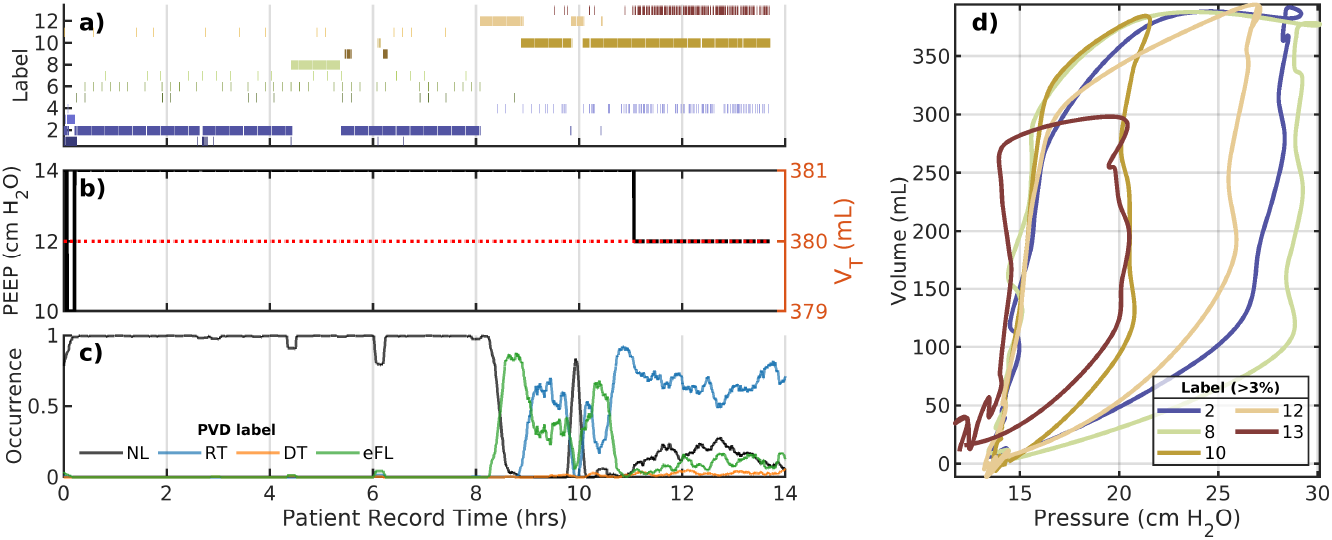
The patient 10 evolution includes only PEEP change. The layout is the same as panels a–d of the previous figure. Under constant ventilator settings, breaths undergo transition several times including intervals of PVD prior to PEEP change around 10.5 hours. A 1-hour long shift from label #2 to #8 occurs around 8 hours during which breaths decrease peak pressure and includes an increase in eFL and RT PVD occurrence. After the PEEP change, LVS behavior is dyssynchronous and primarily centered around the characterization with label #10. Labeling and coloring in this figure do not relate to other figures.

**Figure SI3.**
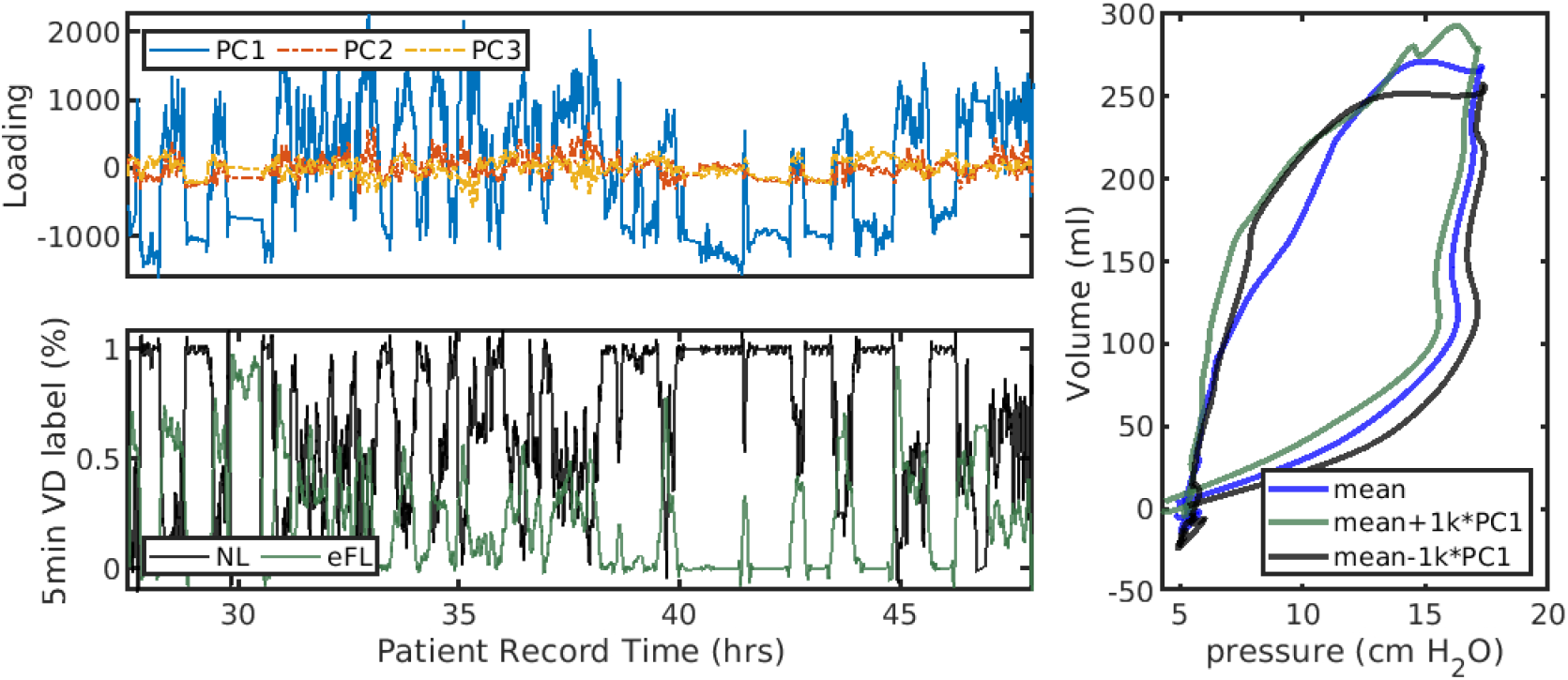
The sign of PC1 loading roughly divides the PVD classes of patient 8, label #2. A threshold for the PC1 loading at zero roughly separates NL and eFL labels by 34%/65% and 85%/14%, respectively, with NL labels strongly associated with negative loadings. The optimal threshold ( ∼ 0.05) offers only subtle improvement. The right panel illustrates low fidelity changes in the cluster median pV loop (blue) when modified by these negative (black, more associated with NL) and positive (green, eFL) loadings. This comparison involves involves linking 10-second properties (representing typically ∼ 3–4 breaths) to breath-wise labels, and some representation errors thus arise from summarizing binary PVD labels over all breaths intersecting a 10-second analysis window. Labeling and coloring in this figure do not relate to other figures.

## SI D Influence of Hyperparameter choices on cohort phenotypes

For each of the 721 individual phenotypes, feature vectors defined by the 5-number summaries of period, PEEP, maxima of volume and pressure, ventilator settings, and estimated parameters of range-normalized waveform were assembled from the population of LVS windows with a given label. Ventilator mode was represented as a vector of percentages of each mode rather than a vector of binary categories. UMAP applied to these cohort feature vectors with the scaled-euclidean metric produced a relatively stable point configuration across various hyper-parameter choices; 12 point neighborhoods (2% of data) with a minimum distance of 1 unit were adopted as values. Identified groups were more sensitive to DBSCAN labeling hyper-parameters. Figure SI 4a shows the possibilities of different groupings based on the search neighborhood size (*ε*). Subsequent results in the main section employ a hyper-parameter choice at the ‘knee-point’^6^ to balance generalizability and specificity. Panel b shows a more specific labeling (*ε* = 2.5) is qualitatively similar to that in the main text.

**Figure SI4.**
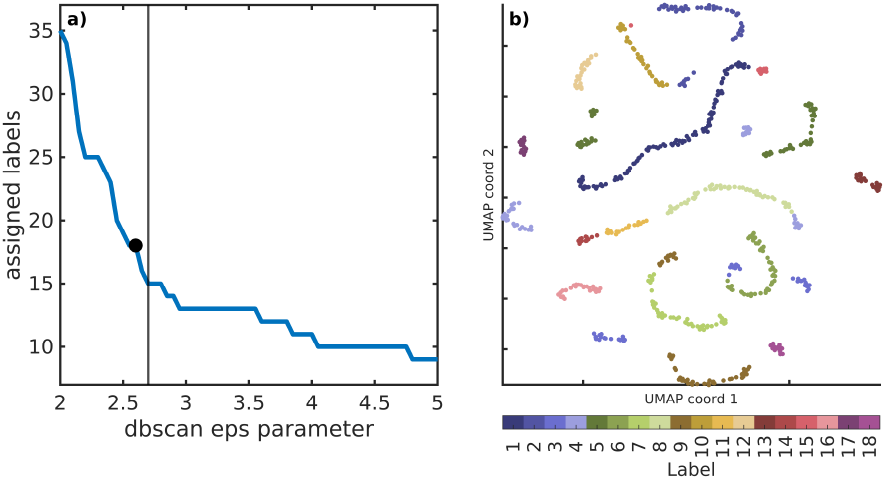
DBSCAN search radius (*ε*) *v*. the number of identified groups. The black line indicates *ε* = 2.7 selected for cohort clustering used in the main text. Choices of *ε* ∈ [2.67, 2.82] yield equivalent results with increasing granularity of groupings for lower values of *ε*. The black dot indicates *ε* = 2.5, whose associated labels are shown (right).

**Table SI.2.** The equivalent of main text Table 2 for the alternate choice of hyper-parameter *ε* = 2.5.

| Label | Total% | $N_{\text{pat}}$ | $N_{\text{pheno}}$ | $p_{\text{min}}$ | $p_{\text{drive}}$ | $V_{\text{max}}$ | dp/dV | MV mode |
| --- | --- | --- | --- | --- | --- | --- | --- | --- |
| 1 | 15.5 | 11 | 101 | 8 | 12.7[4.1] | 7.9[1.3] | 1.5[0.4] | APVCMV |
| 2 | 11.4 | 16 | 52 | 12 | 13.3[3.6] | 6.2[2.1] | 2.3[1.2] | PCMV* |
| 3 | 8.4 | 5 | 37 | 10 | 13.5[1.9] | 6.5[0.3] | 2.1[0.4] | APVCMV* |
| 4 | 7.7 | 8 | 45 | 14 | 12.6[2.9] | 6.2[1.3] | 1.9[0.7] | APVCMV |
| 5 | 6.9 | 11 | 58 | 16 | 13.4[2.3] | 6.0[0.6] | 2.2[0.6] | APVCMV |
| 6 | 6.3 | 7 | 49 | 11 | 16.6[12.5] | 5.9[0.7] | 3.4[2.4] | APVCMV |
| 7 | 6.2 | 6 | 49 | 8 | 11.1[1.4] | 6.6[1.0] | 1.7[0.2] | APVCMV |
| 8 | 6.2 | 23 | 56 | 10 | 12.1[3.7] | 6.3[1.0] | 1.9[0.6] | APVCMV |
| 9 | 6.2 | 10 | 51 | 14 | 21.3[9.5] | 5.6[1.8] | 3.7[3.1] | APVCMV |
| 10 | 4.1 | 12 | 34 | 14 | 12.2[6.9] | 5.9[0.2] | 2.0[1.3] | APVCMV* |
| 11 | 4.0 | 14 | 20 | 5 | 8.9[4.1] | 7.0[2.4] | 1.2[0.8] | APVCMV |
| 12 | 3.7 | 16 | 25 | 12 | 14.3[3.2] | 6.0[0.4] | 2.3[0.9] | PCMV** |
| 13 | 3.4 | 17 | 22 | 11 | 13.1[2.9] | 6.2[1.3] | 2.1[0.5] | APVCMV*** |
| 14 | 3.3 | 11 | 12 | 8 | 9.7[2.7] | 6.8[1.5] | 1.6[0.4] | APVCMV |
| 15 | 2.5 | 8 | 10 | 12 | 15.1[12.5] | 5.9[0.1] | 2.4[2.2] | APVCMV |
| 16 | 1.7 | 11 | 27 | 14 | 13.3[2.9] | 6.0[1.3] | 2.0[0.8] | APVCMV |
| 17 | 1.5 | 9 | 14 | 16 | 15.9[6.0] | 5.9[2.8] | 2.6[2.4] | APVCMV |
| 18 | 1.1 | 5 | 16 | 5 | 10.7[0.2] | 6.5[0.7] | 1.7[0.2] | APVCMV |

